# Long-term survival after invasive pneumococcal disease – a matched cohort study using electronic health records in England

**DOI:** 10.1101/2025.01.31.25321460

**Authors:** Anne Suffel, Fariyo Abdullahi, Eleanor Barry, Jemma Walker, Nick Andrews, Zahin Amin-Chowdhury, Shamez N Ladhani, Daniel Grint, Helen I. McDonald, Ian Douglas, Kathryn E. Mansfield, Edward P.K. Parker

**Author notes:** Joint senior authors (Kathryn E. Mansfield, Edward P.K. Parker). Corresponding author: Anne Suffel.

## Abstract

**Background:** Invasive pneumococcal disease (IPD) is associated with increased long-term mortality, but it is unclear if this can be explained by pre-existing comorbidities that predispose individuals to pneumococcal infection.

**Methods:** We conducted a matched cohort study comparing long-term survival (beyond 120 days after infection) in individuals with IPD and comparators without IPD. Cases were individuals in England aged ≥65+ years with laboratory-confirmed IPD (2012–2019) identified through enhanced national surveillance. Comparators matched on age, sex, and calendar date were drawn from primary care electronic health records in Clinical Practice Research Datalink GOLD. We used Cox regression (stratified by matched set) to compare mortality in people with and without IPD, adjusting for relevant comorbidities, deprivation, and ethnicity.

**Results:** We included 13,401 IPD cases and 67,005 comparators. After adjusting for comorbidities, deprivation, and ethnicity, we found increased all-cause mortality in IPD cases compared to comparators (hazard ratio 3.74, 95% CI: 3.50–3.99). The predicted median survival from adjusted models was 4.7 years (IQR: 2.9-7.4) for IPD cases and >11.9 (IQR : 8.7->11.9) for comparators. This increased mortality was consistent across subgroups defined by age, vaccination history, and comorbidity status (including diabetes, chronic respiratory disease, and chronic heart disease).

**Conclusions:** IPD was associated with increased mortality at least 5 years after infection. These findings demonstrate the long-term consequences of IPD, emphasising the value of IPD prevention and the need for more research into clinical management of IPD survivors. Long-term mortality should be incorporated in cost–effectiveness analyses for pneumococcal vaccines.

**Article summary:** Compared to people without infection, people who survived invasive pneumococcal disease in England had a 3.7-fold increase in mortality up to 10 years after infection. This increased mortality remained after accounting for age, sex, underlying comorbidities, deprivation, and ethnicity.

## Introduction

Invasive pneumococcal disease (IPD) is a severe systemic infection caused by *Streptococcus pneumoniae*, presenting as pneumonia, meningitis, septicaemia, and bacteraemia (1). Annually, in Europe and the USA, there are an estimated 6–10 cases per 100,000 individuals (2,3). Short-term (30-day) case-fatality ratio for IPD ranges from 11 to 30% (1), with higher risk among infants, older adults, and people with underlying health conditions (4). IPD is also associated with increased long-term mortality (5–8). This may reflect a causal effect of IPD on long-term survival; for example, through progression of cardiovascular or chronic kidney disease (9,10). Alternatively, IPD may act as an indicator of underlying frailty and comorbidities (11). The extent to which the poor long-term prognosis after IPD is explained by underlying comorbidities remains unclear as previous studies often compared IPD deaths to standardised population mortality rates (6).

To protect against pneumococcal disease several countries (predominantly high-income) offer the 23-valent pneumococcal polysaccharide vaccine (PPV23) and/or pneumococcal conjugate vaccines (PCV13, PCV15, or PCV20) to older adults and individuals with underlying comorbidities. PPV23 is currently recommended for adults from 65 years of age in the UK (12). Policy decisions regarding the optimal vaccination strategy rely on cost–effectiveness analyses that typically do not account for long-term mortality (13–15). Accurate estimates of the effect of IPD on long-term mortality (accounting for the effect of comorbidities) could support improved cost–effectiveness estimates and promote wider implementation and higher uptake of pneumococcal vaccination.

## Methods

We conducted a cohort study comparing IPD cases identified from national surveillance data to non-IPD comparators using routinely-collected primary care electronic records in England. Comparators were matched to IPD cases based on age, sex, and calendar date.

### Study population

#### IPD cases

Our exposure of interest was IPD (laboratory-confirmed invasive pneumococcal isolate in individuals with a clinical diagnosis of pneumonia, meningitis, or any other invasive infection). We used anonymised data from the UK Health Security Agency (UKHSA) on laboratory-confirmed IPD cases between 1 January 2012 and 31 December 2019. Hospital laboratories routinely report invasive bacterial infections electronically to UHSA and submit invasive pneumococcal isolates to the UKHSA Respiratory and Vaccine Preventable Bacteria Reference Unit for confirmation and serotyping. As part of the national surveillance, all confirmed cases are enhanced followed up via a postal questionnaire to general practitioners (GPs) requesting clinical information on comorbidities and vaccination history (Table 1). (15)

**Table 1.**
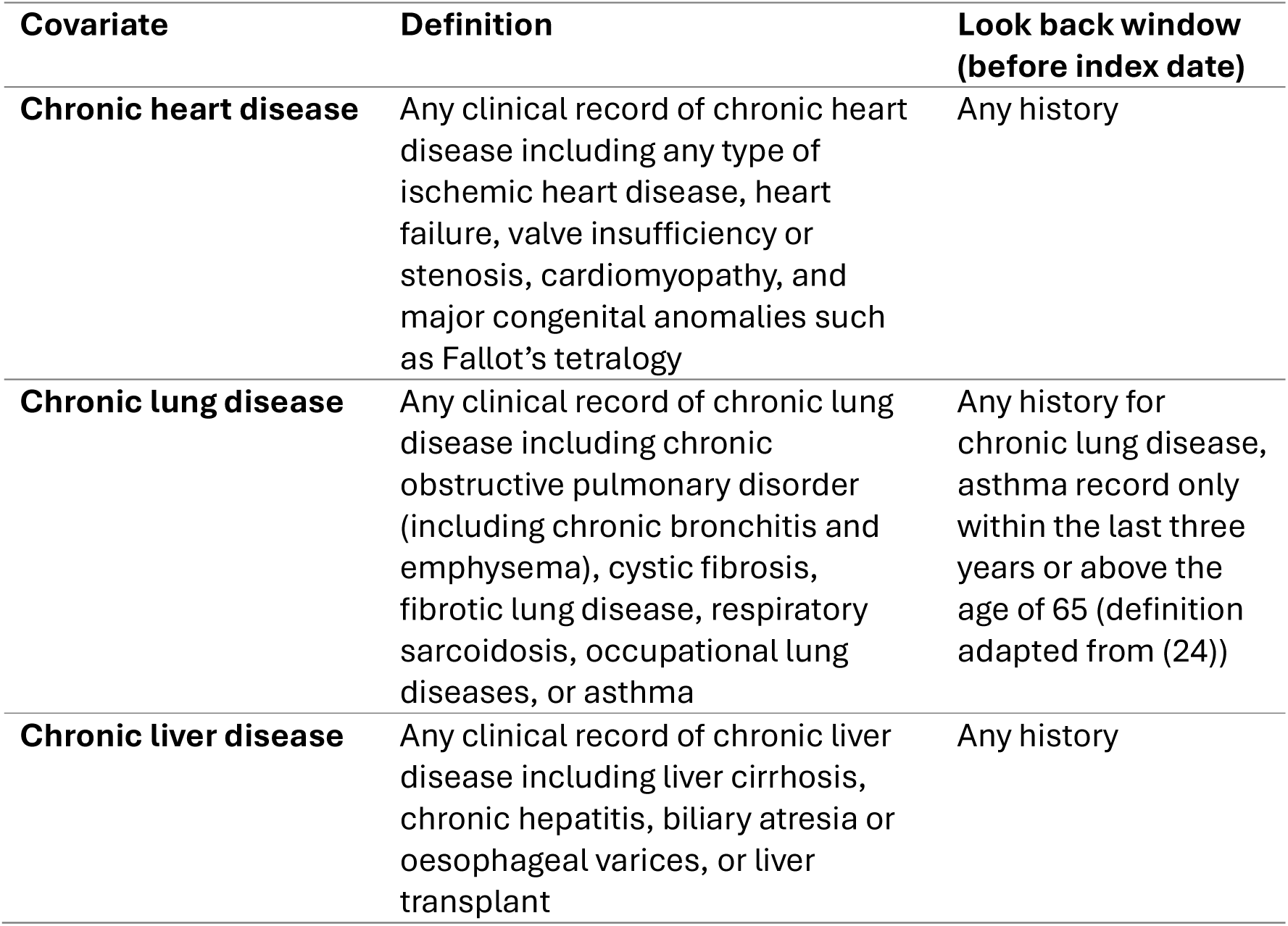
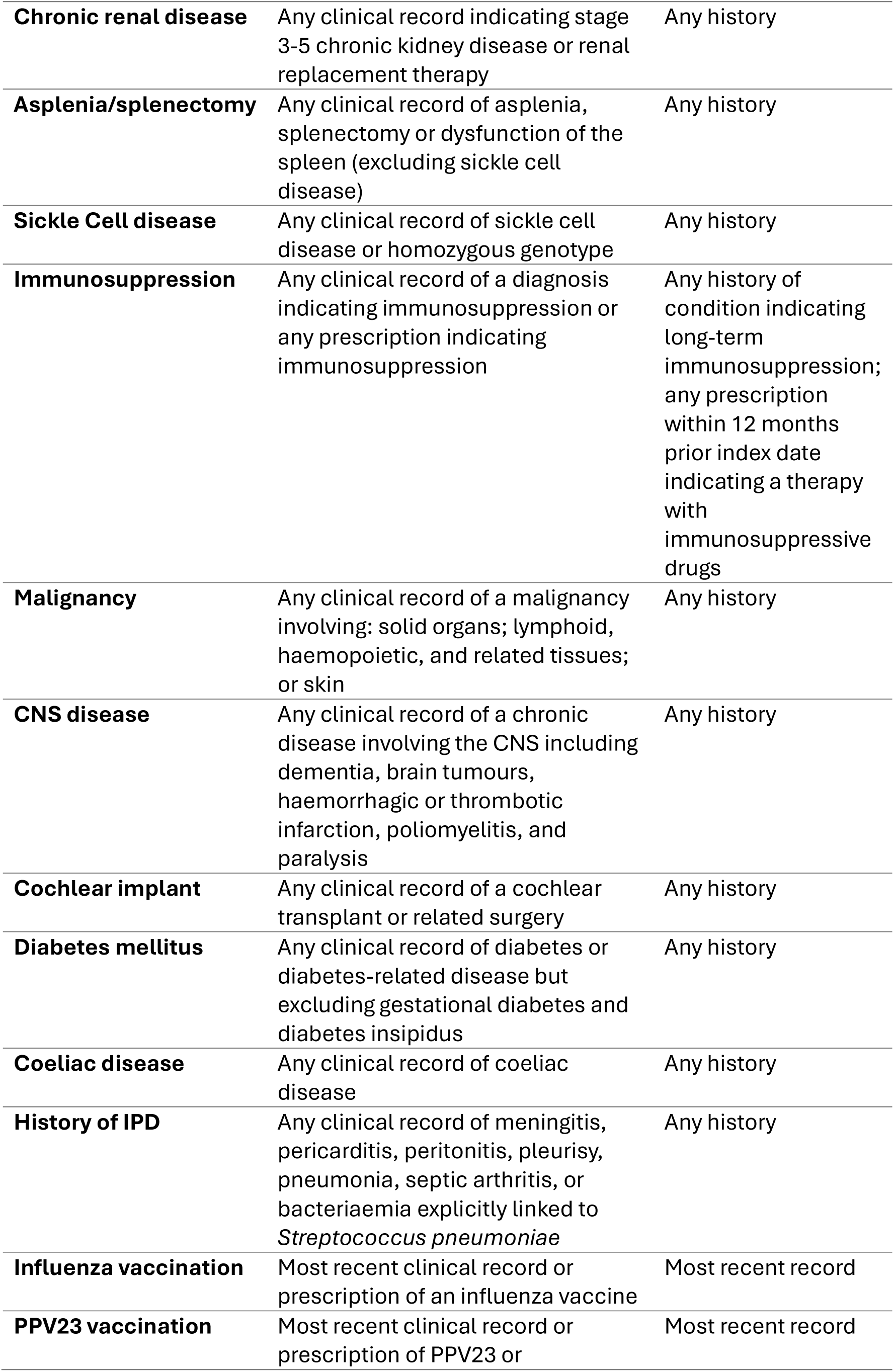
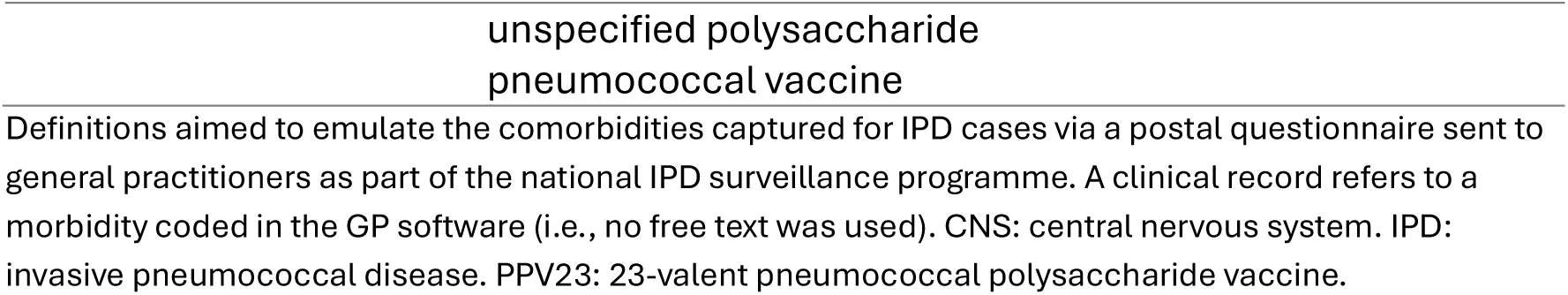
Definition of comorbidities in the non-IPD comparator population.

We included IPD cases aged ≥65 years who survived longer than 120 days after reported infection onset (specimen date). Case data were linked to the Personal Demographics Service (PDS) dataset, which includes date of death in primary care records. Index of multiple deprivation (IMD) quintile was determined based on each individual’s postcode (16). Follow-up started 120 days after IPD infection (index date) and ended at the earliest of death or PDS linkage date (11 April 2024). If cases had more than one eligible IPD episode, we included only the first (interpreting any subsequent episode as a potential mediator on the pathway to increased mortality).

#### Comparators

Non-IPD comparators were derived from the Clinical Practice Research Datalink (CPRD) GOLD primary care database (June 2024 build) (17). The June 2024 CPRD GOLD release holds information on 21,464,230 research quality patients (18). It is broadly representative of the UK population on age, sex, and ethnicity. CPRD GOLD contains information on demographics (sex, year of birth, region), clinical events (symptoms, diagnoses), primary care prescriptions, vaccination, and lifestyle information (e.g., smoking status). We used linked IMD and mortality data from the Office for National Statistics (ONS) where available (19).

Eligible comparators included all individuals who: were at least 65 years old during the surveillance period; registered between 1 January 2012 and 29 April 2020 (corresponding to 120 days after 31 December 2019, i.e., the latest index date of IPD cases); had at least one year of registration; and met CPRD quality-control standards at individual and practice level (20). Comparators entered the cohort on the index date of their matched IPD case (120 days after infection onset). We matched, on year of birth and sex, five comparators, without replacement, to each confirmed IPD case in order of increasing index date. Follow-up ended at the earliest of: death; end of registration with GP practice; last data collection from practice; or end of study period (11 April 2024, aligning with the PDS linkage date of IPD cases).

### Outcome

Our outcome was date of death recorded in primary care records. As linkage to the ONS mortality dataset is considered the gold standard for assessing mortality but was not available for IPD cases (21), we assessed potential outcome misclassification by comparing the date of death in ONS versus primary care records among CPRD comparators with ONS mortality linkage.

### Covariates

For IPD cases, demographic information (year of birth, sex, region of residence, ethnicity [White, Black, South Asian, mixed, other], and IMD quintile) were included in the UKHSA surveillance dataset. Data on the following comorbidities reflecting clinical risk at the time of infection (recorded as “Yes”, “No”, “Unknown”, or blank) were ascertained from the GP questionnaire: chronic heart disease, chronic lung disease, chronic liver disease, chronic renal disease, asplenia or splenectomy, sickle cell disease, immunosuppression or immunosuppressive drug, malignancy, central nervous system (CNS) disease, cochlear implant, diabetes mellitus and coeliac disease (12). PPV23 and influenza vaccination status (and most recent corresponding vaccination dates) were also included.

For comparators, we obtained demographic information (year of birth, sex, region) from primary care records. We defined ethnicity using a validated algorithm (22). We assessed the same comorbidities obtained for IPD cases using relevant clinical (morbidity coded) and prescription records (Table 1). We defined vaccination status using prescription and clinical records (23).

### Statistical analysis

We initially described short-term survival of all IPD cases at 30, 60, 90, and 120 days after infection. We then used Kaplan-Meier curves to estimate long-term survival (from 120 days onwards) for IPD cases and matched non-IPD comparators. We used Cox regression models (stratified by matched set) to estimate hazard ratios (HRs) and 95% confidence intervals (CIs) comparing overall and period-specific (0–1, 0–2, and 0–5 year) mortality in IPD cases (from 120 days post IPD) versus comparators. We estimated a sparse model implicitly accounting for matching variables (age, sex, and calendar date; Model 1), then sequentially adjusted for other covariates as follows: region and IMD quintile (Model 2); additionally adjusting for comorbidities (Model 3; primary analysis); and additionally adjusting for ethnicity (Model 4).

Based on Model 3 (implemented with adjustment for matching variables to enable prediction without stratification index), we estimated survival for both IPD cases and comparators using the Aalen-Johansen approach (25,26). We calculated the median survival time (27) for each individual then the median (IQR) for the predicted survival of the population based on the covariate distribution of the IPD cohort, the comparator cohort, and a standardised IPD cohort (with the same covariate distributions as the comparators). Where the predicted survival probability for an individual was >0.5, the maximum follow-up time was assigned. All analyses were performed in R (Version 4.2.2). Code lists and statistical analysis code are available on GitHub (27).

### Missing Data

We included all eligible IPD cases and comparators in Model 1. Each successive model was restricted to individuals remaining in valid matched sets (cases with at least one comparator after exclusion of individuals with missing data). Individuals with missing region, IMD, or ethnicity were excluded from models adjusting for these. When adjusting for comorbidities (Models 3–4), we included all cases with at least one comorbidity recorded as “Yes” or “No” (indicating at least some engagement with the surveillance questionnaire). If other comorbidities were recorded as “Unknown” or blank, we interpreted this as absence of the comorbidity (but explored this further via sensitivity analysis). For comparators, absence of a medical record for a comorbidity was interpreted as absence of the condition.

### Secondary analyses

We conducted subgroup analyses defined by: age group (65–74, 75–79, 80–89, ≥90 years); comorbidity status (none stated or unknown, cardiovascular disease, respiratory disease, kidney disease, neurological disease, malignancy, diabetes mellitus); and vaccination status (influenza and pneumococcal vaccination history). A series of sensitivity analyses were also performed on Model 3 (Supplementary Table 1).

## Results

We included 13,401 IPD cases and 67,005 matched comparators in the study (Figure 1). Of these, 9422 cases and 47,110 comparators were eligible for the comorbidity-adjusted analyses (hereafter referred to as the primary analysis cohort). Median (IQR) follow-up in the primary analysis cohort was 1,505 days (613–2,205) for IPD cases and 657.5 days (297–4,351) for comparators. Characteristics of IPD cases and comparators are shown in Table 2. Recurrence of IPD during follow-up was observed in 355/13,401 (2.6 %) of cases.

**Figure 1.**
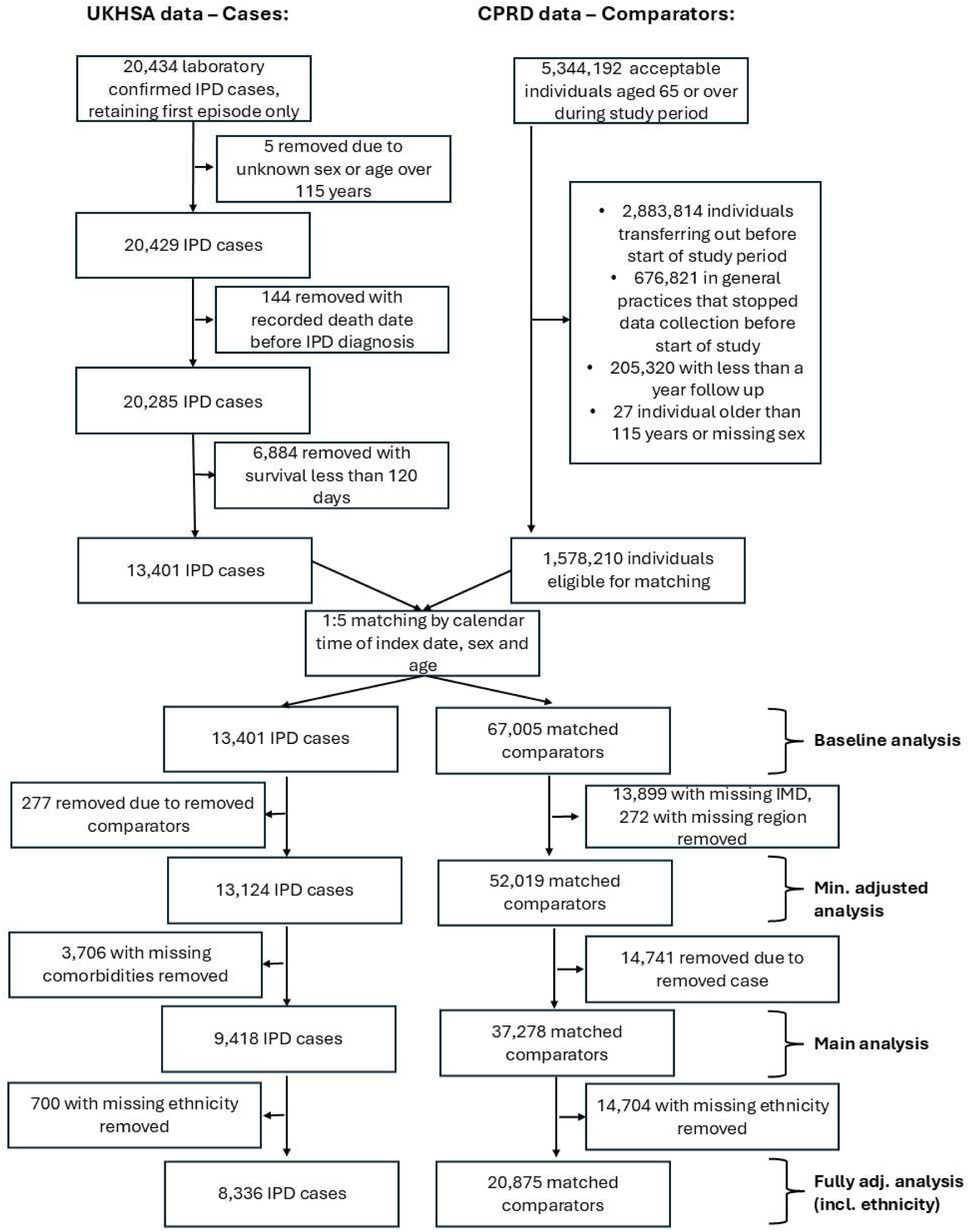
Selection of matched study population. UKHSA: UK Health Security Agency. CPRD: Clinical Practice research datalink; IPD: invasive pneumococcal disease.

**Table 2.**
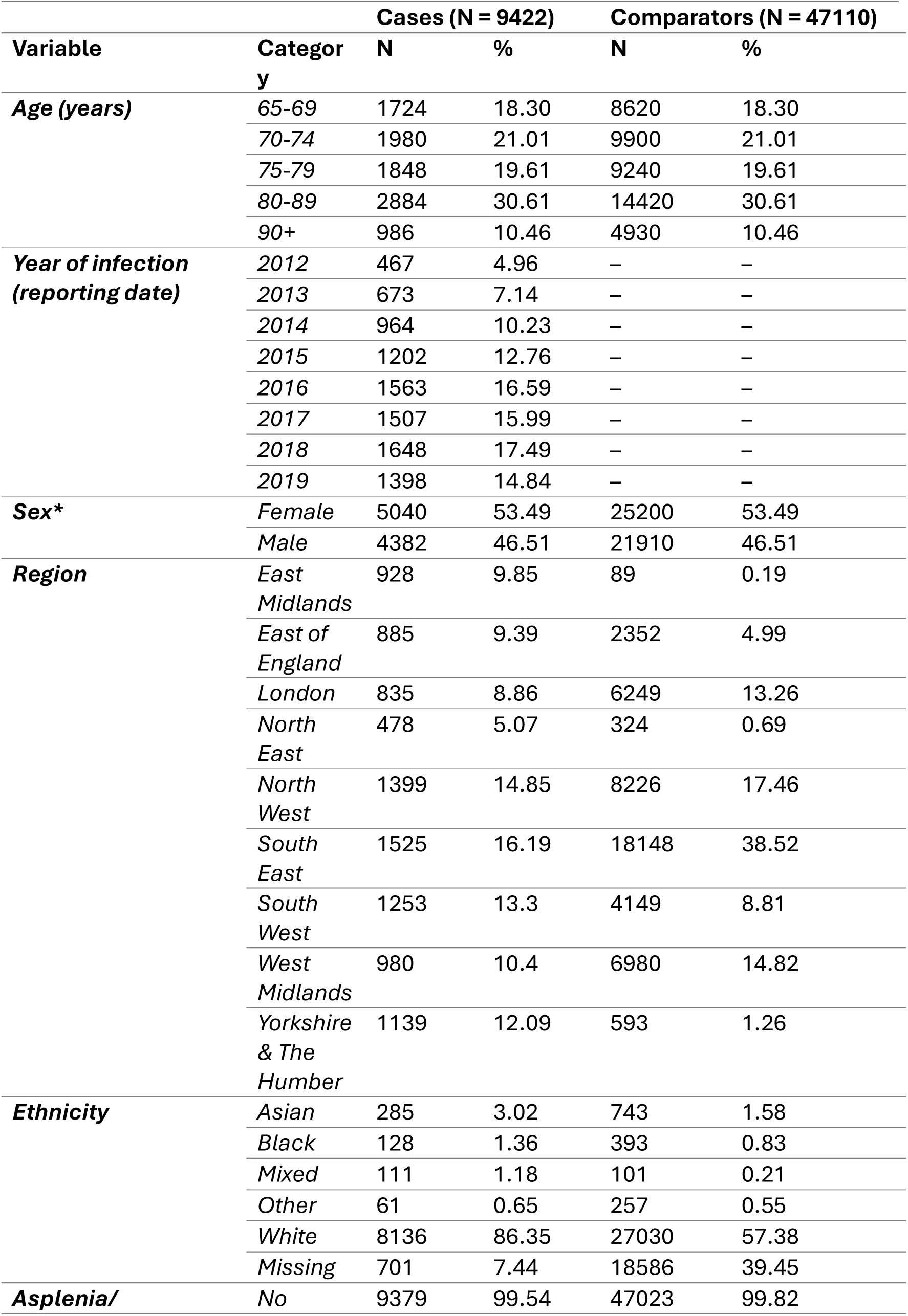
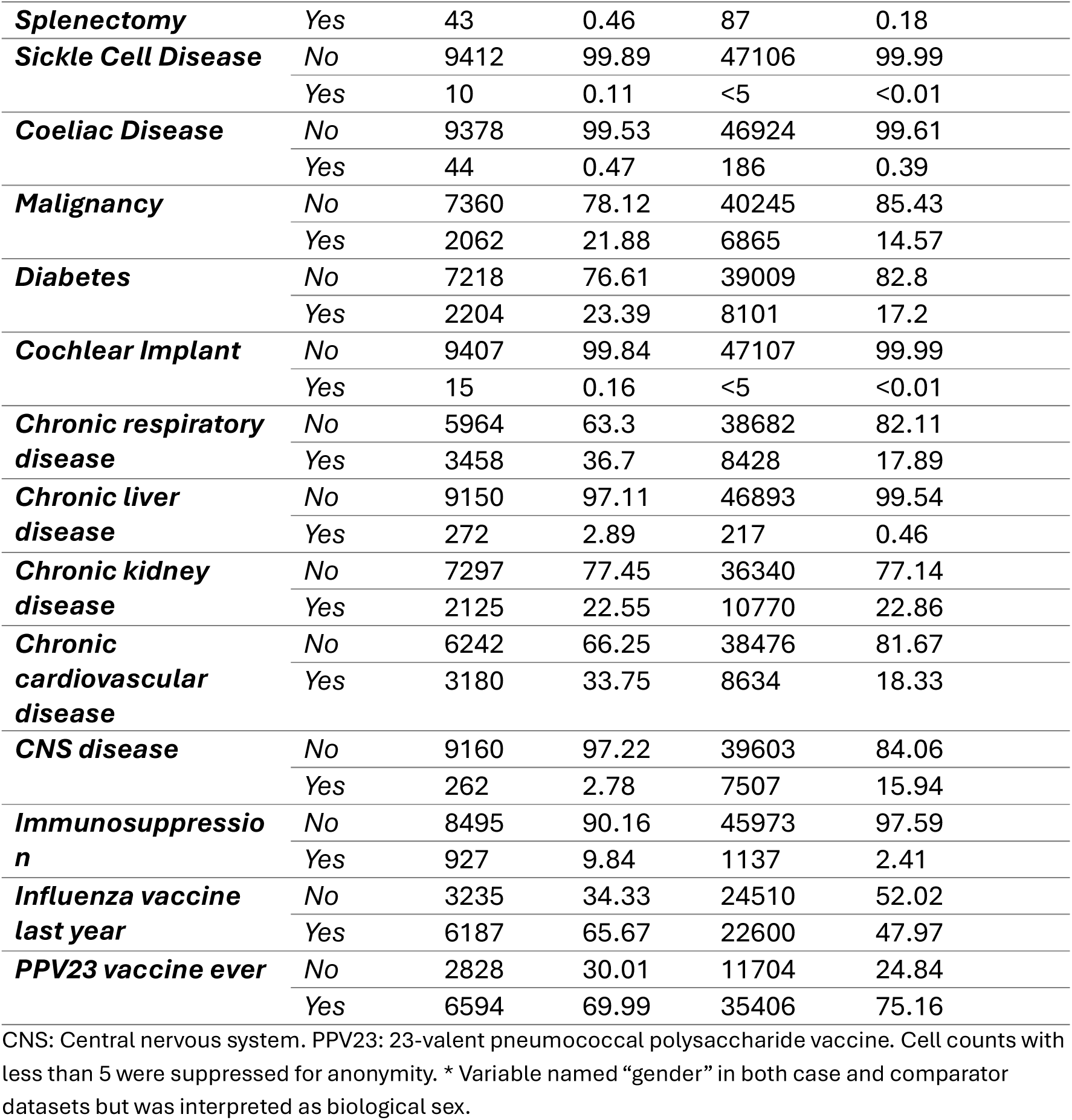
Baseline characteristics of IPD cases who survived to 120 days after infection and their matched comparators.

### Survival in cases vs comparators

5,275/20,285 (26%) IPD cases aged ≥65 years had died within 30 days of infection and 6,884 (34%) died within 120 days (Supplementary Table 2). Among IPD cases who survived past 120 days, mortality was higher than matched non-IPD comparators throughout follow-up (Figure 2). Unadjusted Kaplan-Meier estimates of 1-year survival probability were 83.1% (95% CI: 82.3–83.8%) in IPD cases and 95.4% (95.2–95.6%) in comparators. Unadjusted estimates of 5-year survival were 45.1% (44.1–46.2%) in cases and 79.5% (78.9–80.2%) in comparators.

**Figure 2.**
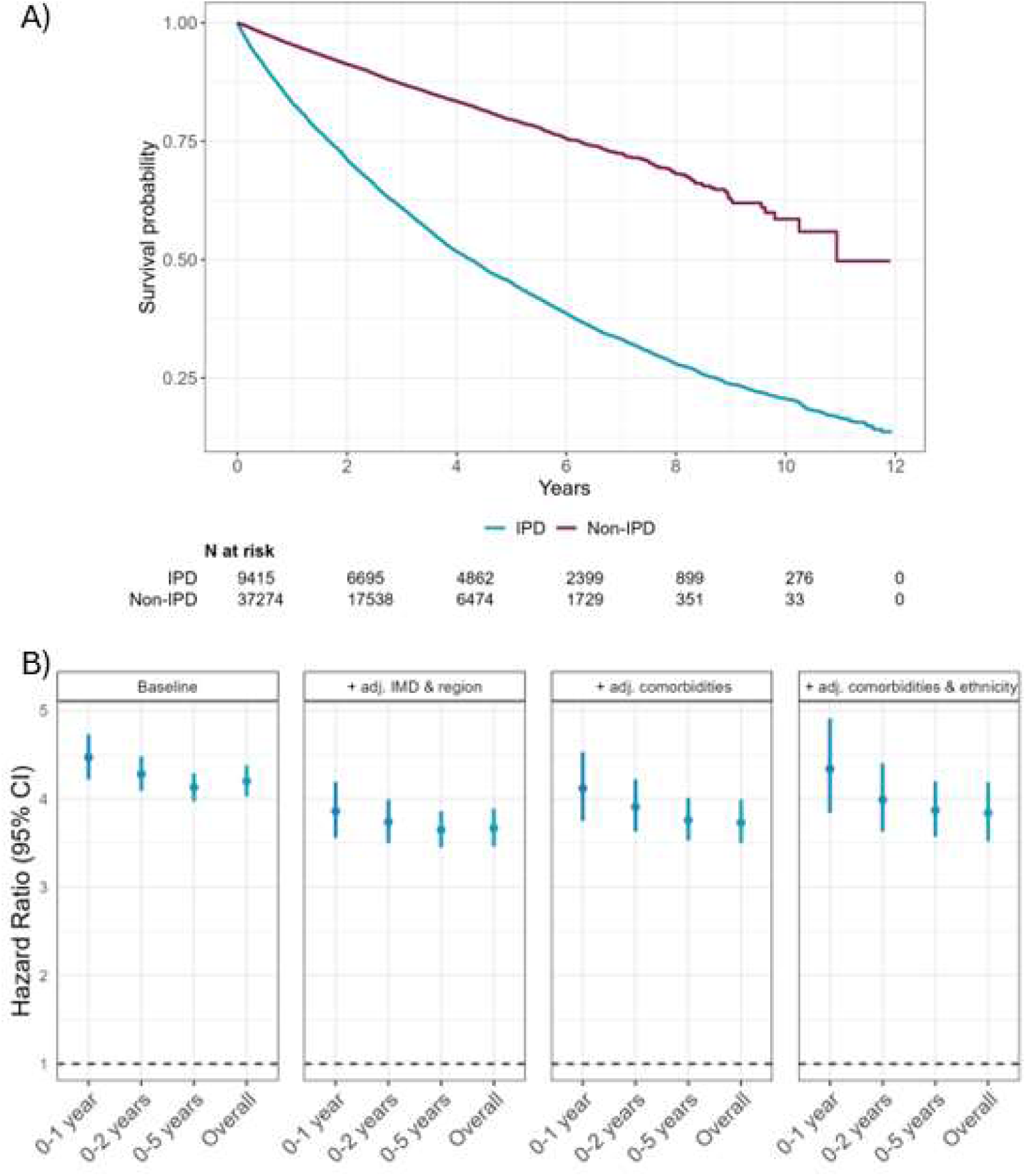
Association between IPD and long-term survival. **A**) Kaplan-Meier curve comparing the survival of individuals with IPD and non-IPD comparators matched by age and sex. For IPD cases, Day 0 (index date) corresponds to 120 days after the recorded date of infection. Kaplan-Meier steps were delayed until 5 events had occurred as a precaution against small number disclosures. **B)**. Hazard ratios (95% CI), estimated using Cox regression stratified by matched set, comparing mortality in IPD cases to non-IPD comparators with sequential adjustment for covariates (region, IMD, comorbidities, ethnicity). See Supplementary Table 5 for full data and supplementary Figure 1 for Kaplan-Meier curves for all cohort definitions. CI: confidence interval; IMD: index of multiple deprivation; IPD: invasive pneumococcal disease.

The HR (95% CI) for all-cause mortality in IPD cases compared to comparators was 4.11 (3.95–4.27) in Model 1 (accounting only for matching variables). When adjusting for region and IMD (Model 2), the HR was 4.14 (3.95–4.35). When further adjusting for pre-existing comorbidities (Model 3), the HR was 3.74 (3.50–3.99), and after further adjusting for ethnicity (Model 4), the HR was 3.84 (3.52–4.18) (see Supplementary Tables 3–5 for baseline characteristics for each sub-selected cohort). Period-specific HRs indicated a slight increase in year-1 excess mortality compared to later periods, although a strong effect of IPD persisted throughout follow-up (Figure 2; Supplementary Table 6).

The median (IQR) predicted survival based on Model 3 (including IMD, region, and comorbidities) was >11.9 (IQR : 8.7->11.9) for non-IPD comparators, 4.7 years (IQR: 2.9- 7.4) for cases, and 5.3 years (3.0-8.7) for cases after demographic and comorbidity standardisation (Figure 3; Supplementary Table 7).

**Figure 3.**
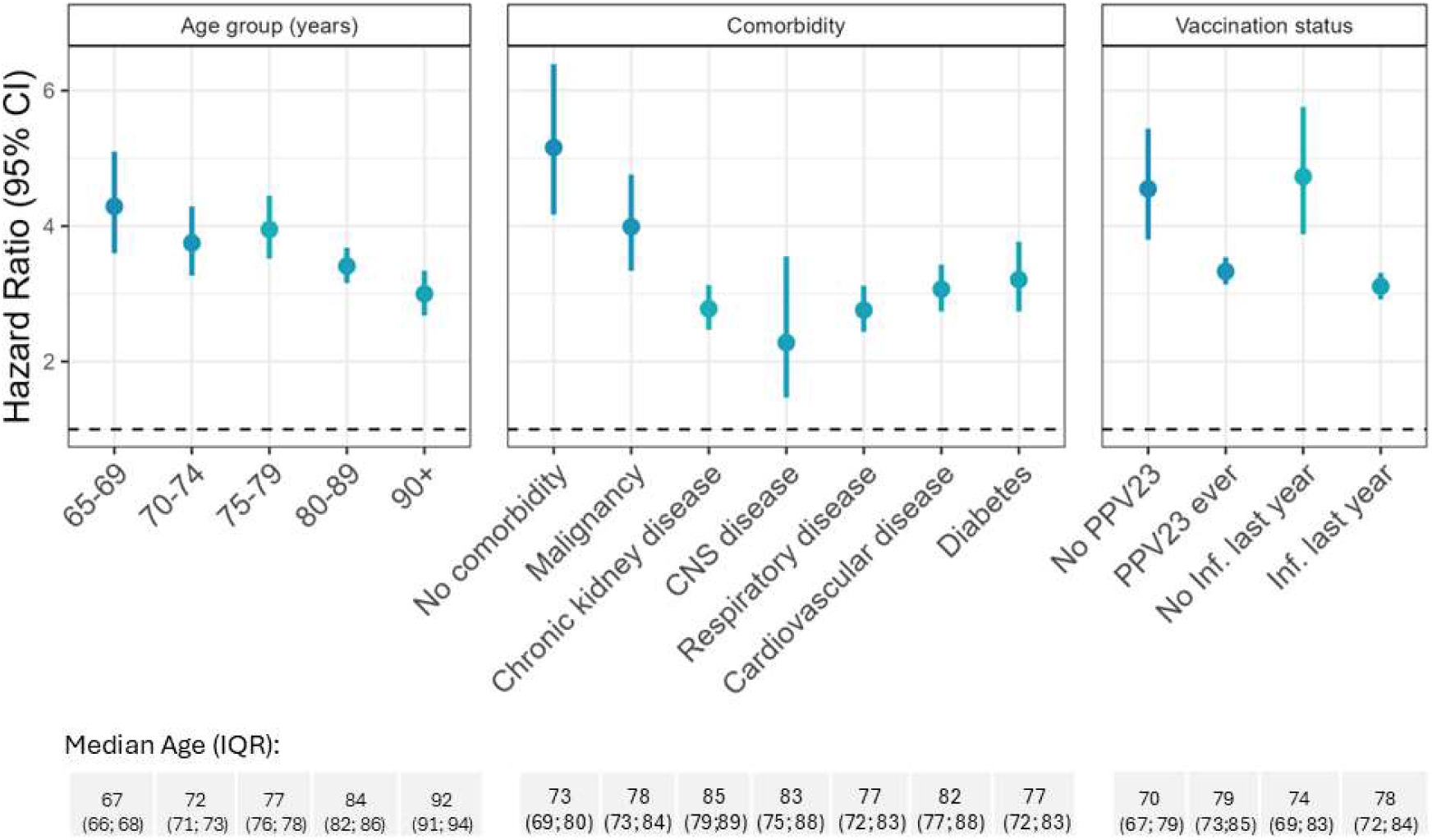
Association between IPD and predicted survival. Predicted median survival and IQR for the overall cohort and stratified by age group. Survival was predicted using the Aalen-Johnson estimator based on models adjusting for region, index of multiple deprivation, underlying comorbidities, and matching variables without stratification. The dashed line indicates the maximum observed follow-up (12 years). For cases, we obtained estimates based on the covariate distribution of the IPD cases and a standardised cohort with the same covariate distributions as the comparator population. See Supplementary Table 7 for full data.

### Secondary analyses

The higher mortality rate in IPD cases compared with comparators was consistent across subgroups defined by age, comorbidity status, and vaccination history (Figure 4; Supplementary Table 8). The lowest HR was observed for individuals between 80–89 years (3.41 [3.16–3.68]) and individuals ≥90 years (3.00 [2.68–3.34])). The corresponding discrepancies in median predicted survival were greater in younger compared with older age groups (Figure 3).

**Figure 4.**
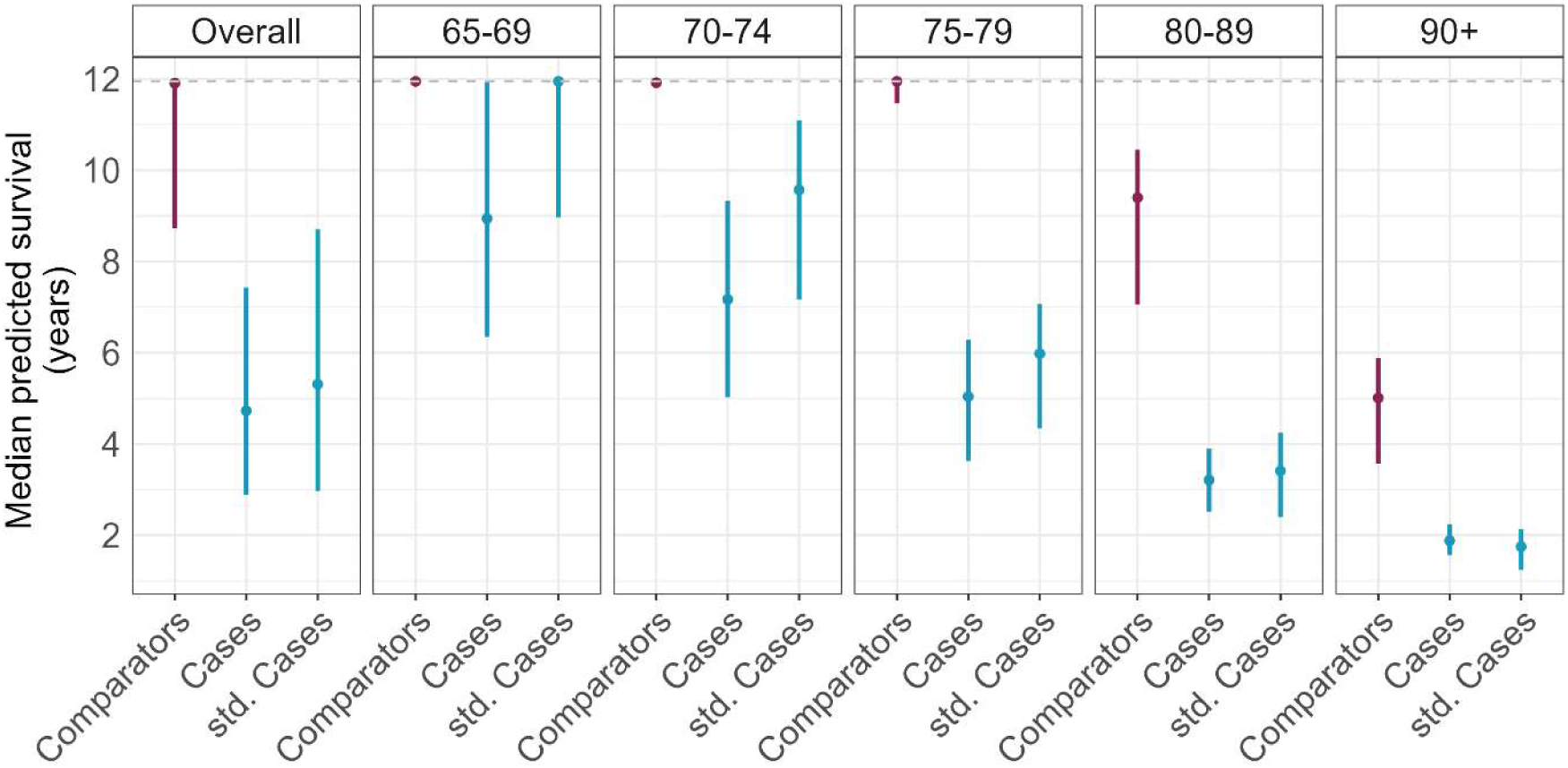
Association between IPD and long-term survival by subgroup. Hazard ratios and 95% CI by age group, underlying comorbidities, and vaccination status. Age of the subpopulations is presented as median (IQR) age in years in the lower panel. See Supplementary Table 8 for full data. Inf: Influenza. IQR: interquartile range; PPV23: pneumococcal polysaccharide vaccine. CNS: central nervous system.

We found a higher HR among individuals with no comorbidities (5.16 [4.17–6.39]) compared to subgroups with diabetes, cardiovascular, respiratory disease, and other common comorbidities (Figure 4). The subgroup of individuals with no comorbidities was younger than subgroups with comorbidities (Figure 4 lower panel). We found a lower HR in individuals with a history of PPV23 vaccine or influenza vaccination in the past year compared with unvaccinated subgroups, though the latter had a notably lower median age.

Results from the following sensitivity analyses were consistent with those of the main analyses: (1) limiting to IPD cases where comorbidities were recorded as “Yes” or “No” (HR 3.72 [3.49–3.97]); (2) excluding comparators with a prior primary care record of IPD (HR 3.75 [3.53–3.99]); (3) limiting follow-up to 31 December 2019 (pre-COVID-19; HR 3.78; [3.52–4.06]); and (4) applying different methods to account for matching (Supplementary Table 7). We observed no indication of selection bias associated with sequential sub-selection of the cohort (Supplementary Table 8).

Combining CPRD data with the linked ONS mortality dataset detected only 215 (2.92%) additional deaths in comparison to using primary care data alone (Supplementary Table 9)

## Discussion

Among IPD cases ≥65 years who survived for 120 days after infection, we documented substantially higher mortality relative to matched non-IPD comparators over a follow-up period spanning up to 12 years. This increased mortality was consistent after adjusting for underlying comorbidities, ethnicity, deprivation, and region, and across subgroups defined by age, comorbidities, and vaccination status. Together, our findings suggest that the increased risk of long-term mortality after IPD cannot be explained by the confounding effect of underlying comorbidities.

Our findings are consistent with previous reports of increased long-term mortality after severe pneumococcal infections. A study of US veterans (392 cases) indicated increased mortality for up to 10 years following pneumococcal pneumonia compared to the general population, with particularly high mortality following severe infections (5). A Norwegian cohort study (372 cases) found increased mortality 10 years after IPD hospital admission, with higher mortality in people with underlying health conditions (6). A Dutch cohort study (228 cases) reported increased mortality up to 3 years after infection for both invasive and non-invasive pneumococcal disease. In contrast to our study, the excess mortality was not apparent among individuals without underlying comorbidities (7). Notably, previous studies have either looked at IPD cases alone or relative to standardised population mortality rates, making it difficult to assess the potentially confounding effect of underlying health conditions.

Our findings support a potential effect of IPD on long-term risk of mortality. This finding is consistent with a growing literature highlighting the role of severe infections in advancing trajectories of frailty, including reduced quality of life, cognitive impairment, and increased mortality (28,29). We lacked data for the cause of death in IPD cases, which could have offered insight into the causal pathway towards increased mortality. However, previous studies have highlighted an increased risk of cardiovascular disease following infection (9,30), potentially linked with persistence of an inflammatory and prothrombotic state (31). Pneumococcal pneumonia has also been linked with increased renal complications including end-stage renal disease (32).

The excess mortality among IPD cases was apparent across all subgroups considered. We found a modestly lower effect size in older compared with younger age groups. This likely reflects the accrual of competing risks of mortality with increasing age, reducing the excess mortality associated with IPD. We also found the excess mortality to be attenuated in subgroups with PPV23 or recent influenza vaccination. This could reflect a protective effect of vaccination on outcomes after IPD (e.g., via a direct effect of PPV23 or a reduction in influenza infections during or after IPD). However, the differences should be interpreted with caution given the older age of vaccinated compared with unvaccinated individuals and the potential for confounding by indication (e.g., higher vaccine uptake in individuals with underlying health conditions that present a competing mortality risk) (33).

Across comorbidities, the effect of IPD was lower in individuals with CNS disease and higher in individuals with malignancy – partially overlapping with findings from a Norwegian study in which cardiovascular disease and malignancy were associated with increased mortality among IPD cases (6). The larger effect size among individuals with no comorbidities may be explained in part by the younger age of this subgroup, although it may also reflect differential ascertainment of comorbidity status in IPD cases (based on GP questionnaire) versus non-IPD comparators (based on primary care records).

Previous cost–effectiveness analyses related to pneumococcal vaccines have focused on short-term mortality (13–15). Our findings support the inclusion of long-term mortality in cost–effectiveness models and provide relative and absolute effect sizes that could be used to parameterise future evaluations, including those relating to new high-valency formulations that incorporate additional serotypes. Notably, 70% of IPD cases and 75% of comparators in this study had been vaccinated with PPV23, despite all individuals in England being eligible for vaccination from the age of 65 years. By highlighting the substantial mortality attributed to IPD, our findings emphasise the long-term harms of severe pneumococcal infection, supporting efforts to promote vaccine uptake. Our study also highlights the need for proactive clinical management of individuals following recovery from IPD to mitigate their higher risk of long-term mortality.

Strengths of our study include the use of nationally representative datasets for IPD cases and comparators. The substantial size of our cohort allowed us to adjust for rare comorbidities. Recruitment from 2012 to 2019 enabled us to estimate effects over a 10- year follow-up period. By using a matched cohort design, it was possible to compare mortality for individuals of the same age, sex, and calendar period, while the detailed investigation of cases via the national IPD surveillance programme enabled us to adjust for relevant comorbidities. Our findings were robust to different modelling strategies and to varying interpretations of missing data in the GP questionnaire.

A key limitation of our study is the use of different data sources used for the IPD cases and comparators, and the differential ascertainment of comorbidities resulting from this. As the comparators could be censored (e.g., by leaving a GP practice) and consequently had a shorter follow-up time than IPD cases, their mortality might have been underestimated, biasing estimates away from the null. GP questionnaires included broad disease categories (e.g., “respiratory disease”). Interpretation of these may have differed across GPs and may not reflect the definitions and look-back periods we applied in non-IPD comparators. Nonetheless, our findings were consistent in analyses restricted to subgroups that are likely to be well defined in both the case and comparator data (e.g., diabetes, for which primary care coding is incentivised by the Quality and Outcomes framework (34)). Other lifestyle and clinical factors such as alcoholism, smoking status, and receipt of medications may also predispose individuals to IPD and higher mortality but could not be accounted for in this study (35). Thus, we cannot exclude the contribution of residual confounding as an explanation for higher mortality observed in IPD cases, though it is unlikely to fully account for the strong effect size observed across models and subgroups. In addition, our study did not consider the effect of pneumococcal serotype, which has previously been shown to influence mortality after IPD (4) and is also a key consideration for vaccine policy decisions. Finally, predicted median survival among comparators exceeded the maximum follow-up time, limiting precise comparison with predicted survival estimates for cases.

In conclusion, IPD impacts health far beyond the initial episode, highlighting the importance of close surveillance and long-term support of patients following IPD. Our findings emphasise the importance of efforts to prevent IPD through encouraging higher vaccination uptake and highlight the relevance of long-term mortality during cost– effectiveness analyses and policy decisions related to pneumococcal vaccination.

## Data Availability

The study uses data from the Clinical Practice Research Datalink (CPRD). CPRD does not allow the sharing of patient-level data. The data specification for the CPRD data set is available at: https://www.cprd.com/doi/cprd-gold-june-2024-dataset. Analysis code and code lists are shared in our online repository under the following link:

## Acknowledgements

This work uses data provided by patients and collected by the NHS as part of their care and support (usemydata.org). This study is based in part on data from the Clinical Practice Research Datalink obtained under licence from the UK Medicines and Healthcare products Regulatory Agency. The data is provided by patients and collected by the NHS as part of their care and support. The interpretation and conclusions contained in this study are those of the author/s alone.

## Authors’ contributions

EPKP, JW, NA, HM, FA, ZAC, SL, DG, and KM conceptualised the study and contributed to the study design. FA processed the IPD case data set at the UKHSA. AS and KM developed the code lists or adapted existing code lists from the Electronic Health Records group at the London School of Hygiene and Tropical Medicine. AS cleaned, analysed, and interpreted the data with input from EPKP, NA, ID, JW, EB and KM. AS wrote the first draft of the manuscript. The manuscript was critically revised by all authors. All authors approved the submission of the article.

## Ethical approval

We received data governance approval from CPRD (protocol number 19_232) and ethical approval from the London School of Hygiene and Tropical Medicine’s research ethics committee (reference number 19198).

