## Supplementary material for "Long-term survival after invasive pneumococcal disease – a matched cohort study using electronic health records in England"

*Supplementary Table 1. Overview of performed sensitivity analyses.*

| **Motivation for sensitivity analysis** | **Analyses performed** |
|---|---|
| Selection Bias due to restricting study population to complete cases | Model 1 before and after excluding individuals with missing IMD and region<br>Model 2 before and after excluding individuals with missing comorbidity information<br>Model 3 before and after excluding individuals with missing ethnicity |
| Ambiguity of comorbidity coding in GP questionnaire | Model 3 was restricting to comorbidities recorded as "Yes" or "No" instead of interpreting absent responses or "Unknown" as an absence of comorbidity (given there was at least record of "Yes" and "No" in the GP questionnaire) |
| Misclassification of exposure | Model 3 was restricted to comparators without any primary care record of an IPD infection (see Table 1 for definition) |
| Matching | To explore the impact of different methods accounting for matching, Model 3 was run with no adjustment for matching, adjustment for the matching variables, and a using a frailty model |
| Disruption of the COVID-19 pandemic (as a cause of mortality and wider healthcare disruption) | Model 3 was run with follow-up ending on the 31 December 2019 |
| Differences in definitions of CNS disease | Instead of using the definition in Table 1, CNS disease in the non-IPD comparators was restricted to medical records of cerebrospinal fluid leak and ventriculoperitoneal shunt as these were given as examples of CNS disease in the GP questionnaire for the IPD cases |

IMD: Index of multiple deprivation. GP: General practice. IPD: invasive pneumococcal disease. CNS: Central nervous system.

*Supplementary Table 2. Description of the short-term IPD mortality.*

| Time of survival | % of individuals (N) |
|---|---|
| < 30 days | 26% (5,275) |
| 30-60 days | 3.68% (746) |
| 60-90 days | 2.44% (495) |
| 90-120 days | 1.81% (368) |
| > 120 days | 44.92% (9,113) |
| Alive | 21.14% (4,288) |

A total of 20,285 cases over 65 years of age from the national IPD surveillance programme were included.

*Supplementary Table 3. Baseline characteristics in full cohort (used for model 1).*

| | | Cases (N = 13,124) | | Comparators (N = 52,019) | |
|---|---|---|---|---|---|
| **Variable** | | **Median** | **IQR** | **Median** | **IQR** |
| *Follow-up in years* | | 3.89 | 1.45-5.95 | 1.88 | 0.85-3.29 |
| *Follow-up in days* | | 1420 | 528-2173 | 685 | 310-1203 |
| **Variable** | **Category** | **N** | **%** | **N** | **%** |
| *Age (years)* | *65-69* | 2362 | 18 | 9368 | 18.01 |
| | *70-74* | 2723 | 20.75 | 10870 | 20.9 |
| | *75-79* | 2554 | 19.46 | 10006 | 19.24 |
| | *80-89* | 4081 | 31.1 | 16258 | 31.25 |
| | *90+* | 1404 | 10.7 | 5517 | 10.61 |
| *Year of infection (reporting date)* | *2012* | 1201 | 9.15 | – | – |
| | *2013* | 1233 | 9.4 | – | – |
| | *2014* | 1288 | 9.81 | – | – |
| | *2015* | 1549 | 11.8 | – | – |
| | *2016* | 1879 | 14.32 | – | – |
| | *2017* | 1885 | 14.36 | – | – |
| | *2018* | 2022 | 15.41 | – | – |
| | *2019* | 2067 | 15.75 | – | – |
| *Gender* | *Female* | 52.68 | 27425 | 52.72 | 52.68 |
| | *Male* | 47.32 | 24594 | 47.28 | 47.32 |
| *Region* | *East Midlands* | 1256 | 9.57 | 115 | 0.22 |
| | *East of England* | 1261 | 9.61 | 2635 | 5.07 |
| | *London* | 1315 | 10.02 | 6132 | 11.79 |
| | *North East* | 661 | 5.04 | 458 | 0.88 |
| | *North West* | 1941 | 14.79 | 10550 | 20.28 |
| | *South East* | 2130 | 16.23 | 19131 | 36.78 |
| | *South West* | 1651 | 12.58 | 5257 | 10.11 |
| | *West Midlands* | 1390 | 10.59 | 6877 | 13.22 |
| | *Yorkshire & The Humber* | 1519 | 11.57 | 864 | 1.66 |

| | | | | | |
|---|---|---|---|---|---|
| *Ethnicity* | *Asian* | 295 | 2.25 | 741 | 1.42 |
| | *Black* | 133 | 1.01 | 414 | 0.8 |
| | *Mixed* | 116 | 0.88 | 109 | 0.21 |
| | *Other* | 65 | 0.5 | 240 | 0.46 |
| | *White* | 8285 | 63.13 | 29746 | 57.18 |
| *Index of multiple deprivation in quintiles* | *1 (least deprived)* | 2821 | 21.49 | 14079 | 27.07 |
| | *2* | 2536 | 19.32 | 11894 | 22.86 |
| | *3* | 2677 | 20.4 | 10634 | 20.44 |
| | *4* | 2632 | 20.05 | 9118 | 17.53 |
| | *5 (most deprived)* | 2458 | 18.73 | 6294 | 12.1 |
| *Asplenia/ Splenectomy* | *Yes* | 43 | 0.33 | 104 | 0.2 |
| | *Unknown* | 581 | 4.43 | - | - |
| | *Missing* | 3563 | 27.15 | - | - |
| *Sickle Cell Disease* | *Yes* | 10 | 0.08 | <5 | <0.01 |
| | *Unknown* | 558 | 4.25 | - | - |
| | *Missing* | 3561 | 27.13 | - | - |
| *Coeliac Disease* | *Yes* | 44 | 0.34 | 213 | 0.41 |
| | *Unknown* | 588 | 4.48 | - | - |
| | *Missing* | 3566 | 27.17 | - | - |
| *Malig-ncy* | *Yes* | 2062 | 15.71 | 7584 | 14.58 |
| | *Unknown* | 490 | 3.73 | - | - |
| | *Missing* | 3557 | 27.1 | - | - |
| *Diabetes* | *Yes* | 2201 | 16.77 | 8840 | 16.99 |
| | *Unknown* | 467 | 3.56 | - | - |
| | *Missing* | 3556 | 27.1 | - | - |
| *Cochlear Implant* | *Yes* | 15 | 0.11 | <5 | <0.01 |
| | *Unknown* | 618 | 4.71 | - | - |
| | *Missing* | 3564 | 27.16 | - | - |
| *Chronic respiratory disease* | *Yes* | 3456 | 26.33 | 9465 | 18.2 |
| | *Unknown* | 423 | 3.22 | - | - |
| | *Missing* | 3549 | 27.04 | - | - |
| *Chronic liver disease* | *Yes* | 272 | 2.07 | 225 | 0.43 |
| | *Unknown* | 604 | 4.6 | - | - |
| | *Missing* | 3563 | 27.15 | - | - |
| *Chronic kidney disease* | *Yes* | 2123 | 16.18 | 12036 | 23.14 |
| | *Unknown* | 490 | 3.73 | - | - |
| | *Missing* | 3556 | 27.1 | - | - |

| | | | | | |
|---|---|---|---|---|---|
| *Chronic cardiovascular disease* | *Yes* | 3178 | 24.22 | 9867 | 18.97 |
| | *Unknown* | 470 | 3.58 | - | - |
| | *Missing* | 3562 | 27.14 | - | - |
| *CNS disease* | *Yes* | 262 | 2 | 8275 | 15.91 |
| | *Unknown* | 608 | 4.63 | - | - |
| | *Missing* | 3564 | 27.16 | - | - |
| *Immunosuppression* | *Yes* | 926 | 7.06 | 1213 | 2.33 |
| | *Unknown* | 661 | 5.04 | - | - |
| | *Missing* | 3577 | 27.26 | - | - |
| *Influenza vaccine last year* | *Yes* | 6288 | 47.91 | 37550 | 72.19 |
| | *Missing* | 5212 | 39.71 | - | - |
| *PPV23 vaccine ever* | *Yes* | 6669 | 50.82 | 39203 | 75.36 |
| | *Unknown* | 84 | 0.64 | - | - |
| | *Missing* | 3521 | 26.83 | - | - |

CNS: Central nervous system. PPV23: 23-valent pneumococcal polysaccharide vaccine. Cell counts with less than 5 were suppressed for anonymity.

*Supplementary Table 4. Baseline characteristics in cohort without missing data on region and index of multiple deprivation (used for model 2).*

| | | Cases (N = 8,336) | | Comparators (N =20,875) | |
|---|---|---|---|---|---|
| **Variable** | | **Median** | **IQR** | **Median** | **IQR** |
| *Follow-up in years* | | 4.13 | 1.72-6.03 | 1.92 | 0.87-3.35 |
| *Follow-up in days* | | 1509 | 627.75-2202.25 | 701 | 316-1223 |
| **Variable** | **Category** | **N** | **%** | **N** | **%** |
| *Age (years)* | *65-69* | 1516 | 18.19 | 3901 | 18.69 |
| | *70-74* | 1781 | 21.37 | 4717 | 22.6 |
| | *75-79* | 1641 | 19.69 | 4155 | 19.9 |
| | *80-89* | 2549 | 30.58 | 6163 | 29.52 |
| | *90+* | 849 | 10.18 | 1939 | 9.29 |
| *Year of infection (reporting date)* | *2012* | 416 | 4.99 | – | – |
| | *2013* | 589 | 7.07 | – | – |
| | *2014* | 835 | 10.02 | – | – |
| | *2015* | 1052 | 12.62 | – | – |
| | *2016* | 1368 | 16.41 | – | – |
| | *2017* | 1338 | 16.05 | – | – |
| | *2018* | 1484 | 17.8 | – | – |
| | *2019* | 1254 | 15.04 | – | – |
| *Gender* | *Female* | 4491 | 53.87 | 11246 | 53.87 |
| | *Male* | 3845 | 46.13 | 9629 | 46.13 |
| *Region* | *East Midlands* | 830 | 9.96 | 27 | 0.13 |
| | *East of England* | 768 | 9.21 | 709 | 3.4 |
| | *London* | 729 | 8.75 | 3234 | 15.49 |
| | *North East* | 431 | 5.17 | 135 | 0.65 |
| | *North West* | 1230 | 14.76 | 4571 | 21.9 |
| | *South East* | 1326 | 15.91 | 7081 | 33.92 |
| | *South West* | 1109 | 13.3 | 1910 | 9.15 |
| | *West Midlands* | 877 | 10.52 | 2750 | 13.17 |
| | *Yorkshire & The* | 1036 | 12.43 | 458 | 2.19 |

| | | | | | |
|---|---|---|---|---|---|
| | *Humber* | | | | |
| *Ethnicity* | *Asian* | 271 | 3.25 | 492 | 2.36 |
| | *Black* | 123 | 1.48 | 276 | 1.32 |
| | *Mixed* | 102 | 1.22 | 77 | 0.37 |
| | *Other* | 59 | 0.71 | 162 | 0.78 |
| | *White* | 7781 | 93.34 | 19868 | 95.18 |
| *Index of multiple deprivation in quintiles* | *1 (least deprived)* | 1789 | 21.46 | 5372 | 25.73 |
| | *2* | 1605 | 19.25 | 4827 | 23.12 |
| | *3* | 1683 | 20.19 | 4188 | 20.06 |
| | *4* | 1679 | 20.14 | 3996 | 19.14 |
| | *5 (most deprived)* | 1580 | 18.95 | 2492 | 11.94 |
| *Asplenia/ Splenectomy* | *Yes* | 38 | 0.46 | 45 | 0.22 |
| *Sickle Cell Disease* | *Yes* | 8 | 0.1 | <5 | <0.01 |
| *Coeliac Disease* | *Yes* | 33 | 0.4 | 92 | 0.44 |
| *Malignancy* | *Yes* | 1806 | 21.67 | 3129 | 14.99 |
| *Diabetes* | *Yes* | 1973 | 23.67 | 3688 | 17.67 |
| *Cochlear Implant* | *Yes* | 13 | 0.16 | <5 | <0.01 |
| *Chronic respiratory disease* | *Yes* | 3104 | 37.24 | 4000 | 19.16 |
| *Chronic liver disease* | *Yes* | 247 | 2.96 | 104 | 0.5 |
| *Chronic kidney disease* | *Yes* | 1897 | 22.76 | 4765 | 22.83 |
| *Chronic cardiovascular disease* | *Yes* | 2817 | 33.79 | 3949 | 18.92 |
| *CNS disease* | *Yes* | 235 | 2.82 | 3317 | 15.89 |
| *Immunosuppression* | *Yes* | 829 | 9.94 | 491 | 2.35 |
| *Influenza vaccine last year* | *Yes* | 5522 | 66.24 | 15543 | 74.46 |
| *PPV23 vaccine ever* | *Yes* | 5871 | 70.43 | 16331 | 78.23 |

CNS: Central nervous system. PPV23: 23-valent pneumococcal polysaccharide vaccine. Cell counts with less than 5 were suppressed for anonymity.

*Supplementary Table 5. Baseline characteristics in cohort without missing ethnicity data (used for model 4).*

| | | Cases (N = 8,336) | | Comparators (N =20,875) | |
|---|---|---|---|---|---|
| **Variable** | | **Median** | **IQR** | **Median** | **IQR** |
| *Follow-up in years* | | 4.13 | 1.72-6.03 | 1.92 | 0.87-3.35 |
| *Follow-up in days* | | 1509 | 627.75-2202.25 | 701 | 316-1223 |
| **Variable** | **Category** | **N** | **%** | **N** | **%** |
| *Age (years)* | *65-69* | 1516 | 18.19 | 3901 | 18.69 |
| | *70-74* | 1781 | 21.37 | 4717 | 22.6 |
| | *75-79* | 1641 | 19.69 | 4155 | 19.9 |
| | *80-89* | 2549 | 30.58 | 6163 | 29.52 |
| | *90+* | 849 | 10.18 | 1939 | 9.29 |
| *Year of infection (reporting date)* | *2012* | 416 | 4.99 | – | – |
| | *2013* | 589 | 7.07 | – | – |
| | *2014* | 835 | 10.02 | – | – |
| | *2015* | 1052 | 12.62 | – | – |
| | *2016* | 1368 | 16.41 | – | – |
| | *2017* | 1338 | 16.05 | – | – |
| | *2018* | 1484 | 17.8 | – | – |
| | *2019* | 1254 | 15.04 | – | – |
| *Gender* | *Female* | 4491 | 53.87 | 11246 | 53.87 |
| | *Male* | 3845 | 46.13 | 9629 | 46.13 |
| *Region* | *East Midlands* | 830 | 9.96 | 27 | 0.13 |
| | *East of England* | 768 | 9.21 | 709 | 3.4 |
| | *London* | 729 | 8.75 | 3234 | 15.49 |
| | *North East* | 431 | 5.17 | 135 | 0.65 |
| | *North West* | 1230 | 14.76 | 4571 | 21.9 |
| | *South East* | 1326 | 15.91 | 7081 | 33.92 |
| | *South West* | 1109 | 13.3 | 1910 | 9.15 |
| | *West Midlands* | 877 | 10.52 | 2750 | 13.17 |
| | *Yorkshire & The* | 1036 | 12.43 | 458 | 2.19 |

| | | | | | |
|---|---|---|---|---|---|
| | *Humber* | | | | |
| *Ethnicity* | *Asian* | 271 | 3.25 | 492 | 2.36 |
| | *Black* | 123 | 1.48 | 276 | 1.32 |
| | *Mixed* | 102 | 1.22 | 77 | 0.37 |
| | *Other* | 59 | 0.71 | 162 | 0.78 |
| | *White* | 7781 | 93.34 | 19868 | 95.18 |
| *Index of multiple deprivation in quintiles* | *1 (least deprived)* | 1789 | 21.46 | 5372 | 25.73 |
| | *2* | 1605 | 19.25 | 4827 | 23.12 |
| | *3* | 1683 | 20.19 | 4188 | 20.06 |
| | *4* | 1679 | 20.14 | 3996 | 19.14 |
| | *5 (most deprived)* | 1580 | 18.95 | 2492 | 11.94 |
| *Asplenia/ Splenectomy* | *Yes* | 38 | 0.46 | 45 | 0.22 |
| *Sickle Cell Disease* | *Yes* | 8 | 0.1 | <5 | <0.01 |
| *Coeliac Disease* | *Yes* | 33 | 0.4 | 92 | 0.44 |
| *Malignancy* | *Yes* | 1806 | 21.67 | 3129 | 14.99 |
| *Diabetes* | *Yes* | 1973 | 23.67 | 3688 | 17.67 |
| *Cochlear Implant* | *Yes* | 13 | 0.16 | <5 | <0.01 |
| *Chronic respiratory disease* | *Yes* | 3104 | 37.24 | 4000 | 19.16 |
| *Chronic liver disease* | *Yes* | 247 | 2.96 | 104 | 0.5 |
| *Chronic kidney disease* | *Yes* | 1897 | 22.76 | 4765 | 22.83 |
| *Chronic cardiovascular disease* | *Yes* | 2817 | 33.79 | 3949 | 18.92 |
| *CNS disease* | *Yes* | 235 | 2.82 | 3317 | 15.89 |
| *Immunosuppression* | *Yes* | 829 | 9.94 | 491 | 2.35 |
| *Influenza vaccine last year* | *Yes* | 5522 | 66.24 | 15543 | 74.46 |
| *PPV23 vaccine ever* | *Yes* | 5871 | 70.43 | 16331 | 78.23 |

CNS: Central nervous system. PPV23: 23-valent pneumococcal polysaccharide vaccine. Cell counts with less than 5 were suppressed for anonymity.

*Supplementary Table 6. Primary analysis with overall and period-specific hazard ratios for the association between IPD and mortality.*

| Model | Follow-up period | N | N events | HR (95% CI) |
|---|---|---|---|---|
| Baseline (model 1) | 0-1 year | 80,406 | 5,199 | 4.47 (4.22-4.73) |
| | 0-2 years | 80,406 | 8,720 | 4.28 (4.09-4.48) |
| | 0-5 years | 80,406 | 14,240 | 4.13 (3.97-4.29) |
| | Overall | 80,406 | 16,315 | 4.11 (3.95-4.27) |
| Plus adjustment for IMD and region (model 2) | 0-1 year | 65,143 | 4,592 | 4.47 (4.16-4.80) |
| | 0-2 years | 65,143 | 7,709 | 4.30 (4.06-4.56) |
| | 0-5 years | 65,143 | 12,749 | 4.14 (3.94-4.35) |
| | Overall | 65,143 | 14,741 | 4.14 (3.95-4.35) |
| Plus adjustment for comorbidities (model 3) | 0-1 year | 46,696 | 3,075 | 4.06 (3.67-4.49) |
| | 0-2 years | 46,696 | 5,162 | 3.91 (3.61-4.24) |
| | 0-5 years | 46,696 | 8,803 | 3.73 (3.49-4.00) |
| | Overall | 46,696 | 10,252 | 3.74 (3.50-3.99) |
| Plus adjustment for ethnicity (model 4) | 0-1 year | 29,211 | 2,184 | 4.28 (3.75-4.90) |
| | 0-2 years | 29,211 | 3,706 | 3.98 (3.59-4.42) |
| | 0-5 years | 29,211 | 6,559 | 3.86 (3.53-4.21) |
| | Overall | 29,211 | 7,772 | 3.84 (3.52-4.18) |

CI: confidence interval; HR: hazard ratio; IPD: invasive pneumococcal disease.

*Supplementary Table 7. Predicted survival times for comparators, cases and cases standardized to the demographics of the control group.*

| Cohort | Group | Median of the predicted restricted mean survival time (IQR) |
|---|---|---|
| 65-69 | Comparators | 11.22 (10.81-11.35) |
| | Cases | 8.00 (6.50-9.23) |
| | std. Cases | 9.23 (8.03-9.65) |
| 70-74 | Comparators | 10.67 (10.18-10.91) |
| | Cases | 6.97 (5.7-8.01) |
| | std. Cases | 8.07 (6.96-8.70) |
| 75-79 | Comparators | 9.86 (9.04-10.23) |
| | Cases | 5.42 (4.36-6.42) |
| | std. Cases | 6.20 (4.88-6.92) |
| 80-89 | Comparators | 8.00 (6.86-8.68) |
| | Cases | 3.93 (3.22-4.64) |
| | std. Cases | 4.13 (3.06-4.93) |
| 90+ | Comparators | 5.68 (4.51-6.41) |
| | Cases | 2.56 (2.14-3.02) |
| | std. Cases | 2.38 (1.73-2.89) |
| Overall | Comparators | 10.75 (9.46-11.95) |
| | Cases | 7.27 (5.69-11.95) |
| | std. Cases | 9.05 (6.37-11.95) |

Survival was predicted using the Aalen-Johnson estimator based on models adjusting for region, index of multiple deprivation, and underlying comorbidities (model 3). For cases, we obtained estimates based on the covariate distribution of the IPD cases and a standardized cohort with the same covariate distributions as the comparator population. CI: confidence interval; IQR: interquartile range.

***Supplementary Table 8***. *Sensitivity analyses.*

| Type of sensitivity analysis | Cohort | Model | N | N events | HR (95% CI) |
|---|---|---|---|---|---|
| Selection bias after restricting to no missing region and IMD | Full cohort | Model 1 (baseline) | 80406 | 16315 | 4.11 (3.95-4.27) |
| | Cohort with complete data on region and IMD | Model 1 (baseline) | 65143 | 14741 | 4.2 (4.03-4.38) |
| Selection bias after restricting to cases with valid comorbidity recording | Cohort with complete data on region and IMD | Model 2 (IMD and region) | 65143 | 14741 | 4.14 (3.95-4.35) |
| | Cohort with valid comorbidity recording (primary analysis cohort) | Model 2 (IMD and region) | 46696 | 10252 | 3.67 (3.46-3.89) |
| Selection bias after restricting to individuals with no missing ethnicity data | Cohort with valid comorbidity recording (primary analysis cohort) | Model 3 (comorbidities) | 46696 | 10252 | 3.74 (3.5-3.99) |
| | Cohort with complete ethnicity data | Model 3 (comorbidities) | 29211 | 7772 | 3.82 (3.5-4.16) |
| Removing IPD cases in comparator population | Primary analysis cohort | Model 3 (comorbidities) | 46696 | 10252 | 3.74 (3.5-3.99) |
| | IPD exclusion in comparators | Model 3 (comorbidities) | 46531 | 10221 | 3.76 (3.52-4.02) |
| Including or excluding unknown values in GP questionnaire | Primary analysis cohort | Model 3 (comorbidities) | 46696 | 10252 | 3.74 (3.50-3.99) |
| | Cohort with complete recording of comorbidities in IPD cases | Model 3 (comorbidities) | 41822 | 9234 | 3.70 (3.45-3.97) |
| Limiting follow-up to 31 December 2019 | Primary analysis cohort | Model 3 (comorbidities) | 46696 | 10252 | 3.74 (3.50-3.99) |
| | Primary analysis cohort restricting follow-up to 31 December 2019 | Model 3 (comorbidities) | 46696 | 6147 | 3.74 (3.46-4.04) |
| Different methods to adjust for the matching | Primary analysis cohort | Model 3 stratified by matching | 46696 | 10252 | 3.74 (3.5-3.99) |
| | Primary analysis cohort | Model 3 with no adjustment for matching | 46696 | 10252 | 3.44 (3.28-3.61) |
| | Primary analysis cohort | Model 3 with adjustment for | 46696 | 10252 | 3.55 (3.38-3.72) |

| | | | | | |
|---|---|---|---|---|---|
| | | matching variables | | | |
| | Primary analysis cohort | Model 3 with frailty term based on matching ID | 46696 | 10252 | 3.66 (3.48-3.84) |

CI: confidence interval; HR: hazard ratio; IMD: index of multiple deprivation; IPD: invasive pneumococcal disease.

***Supplementary Table 9.*** *Difference in death recording between CPRD GOLD and ONS mortality linkage among comparators eligible for ONS linkage (N = 50,201)*

| | |
|---|---|
| *Overall number of deaths recorded by dataset* | |
| Number of deaths recorded in CPRD | 5,779 |
| Number of deaths recorded in ONS prior to censoring | 5,207 |
| Number of deaths recorded in either CPRD or ONS | 5,5995 |
| Number of deaths recorded in both CPRD and ONS | 4,991 |
| Number of deaths recorded in ONS only | 215 |
| Number of deaths recorded in CPRD only | 641 |
| *Differences in date recording of death date where recorded in both CPRD and ONS* | |
| Number (%) of deaths recorded on the same day between both datasets for individuals eligible for ONS linkage | 3,839 (74.70%) |
| Median number (IQR) difference in days between ONS mortality linkage death date and CPRD death date in individuals eligible for ONS linkage | 0 (0-1) |
| Number (%) of individuals with more than a week difference of recorded death date between ONS mortality linkage and CRPD | 576 (9.61%) |

CPRD: Clinical Practice Research Datalink. ONS: Office for National Statistics. IQR: Interquartile range. [1]The number of deaths recorded in either of the datasets (N = 7,417) was used as denominator. [2]The number of deaths in individuals eligible for linkage was used as a denominator (N= 5,995).

***Supplementary Table 10.*** *Secondary analyses stratified by vaccination status, age group and underlying comorbidities.*

| Subgroup | Stratified by | N | N events | HR (95% CI) |
|---|---|---|---|---|
| Vaccination status | PPV23 ever | 26,422 | 6,988 | 3.33 (3.14-3.54) |
| | No PPV23 | 5,044 | 1,210 | 4.55 (3.8-5.44) |
| | Inf. last year | 23,893 | 6,386 | 3.11 (2.92-3.31) |
| | No Inf. last year | 3,546 | 1,049 | 4.73 (3.88-5.76) |
| Age group | 65–79 years | 8,561 | 996 | 4.29 (3.6-5.10) |
| | 70–74 years | 9,872 | 1,450 | 3.75 (3.27-4.29) |
| | 75–79 years | 9,056 | 1,835 | 3.95 (3.52-4.45) |
| | 80–89 years | 14,360 | 4,102 | 3.41 (3.16-3.68) |
| | 90+ years | 4,847 | 1,869 | 3.00 (2.68-3.34) |
| Comorbidity | No comorbidity | 4,523 | 818 | 5.16 (4.17-6.39) |
| | Diabetes | 2,768 | 1,056 | 3.21 (2.74-3.77) |
| | Cardiovascular Disease | 4,371 | 1,953 | 3.07 (2.74-3.43) |
| | Respiratory Disease | 4,418 | 1,790 | 2.76 (2.44-3.12) |
| | CNS Disease | 316 | 144 | 2.28 (1.47-3.55) |
| | Chronic Kidney Disease | 3,820 | 1,678 | 2.78 (2.47-3.13) |
| | Malignancy | 2,176 | 923 | 3.99 (3.34-4.76) |

IPD-associated HRs were obtained from models adjusting for region, index of multiple deprivation, and underlying comorbidities (model 3). CI: confidence interval; CNS: central nervous system; HR: hazard ratio.

### *Supplementary Figure 1.*

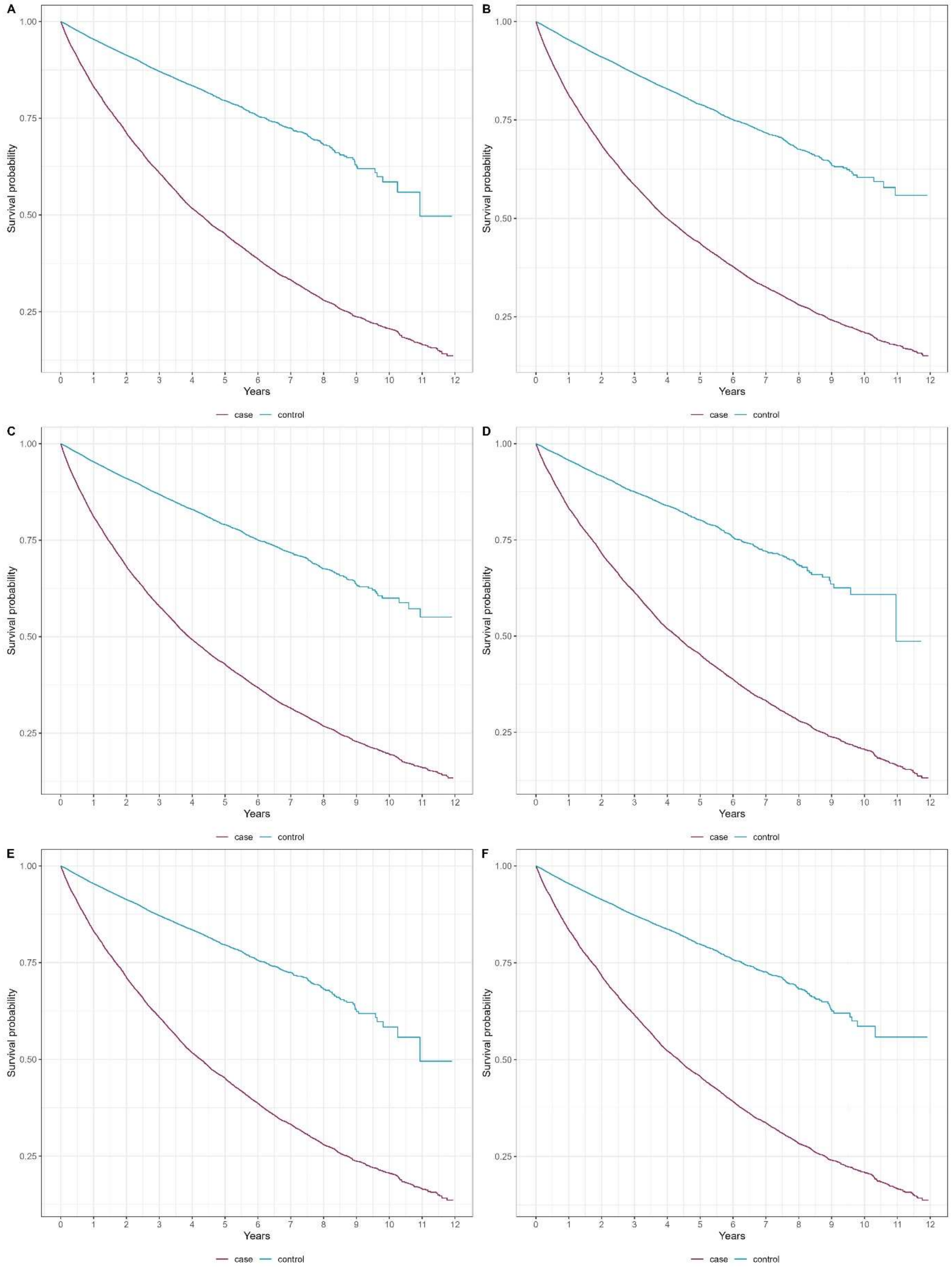


**Figure S1.** Kaplan-Meier curve comparing the survival of individuals with IPD and non-IPD comparators matched by age, gender, and calendar date. For IPD cases, Day 0 (index date) corresponds 120 days after the recorded date of infection. Kaplan-Meier steps were delayed until 5 events had occurred as a precaution against small number disclosures. A) compares the survival for the main study cohort (individuals with missing deprivation, region and comorbidity data removed), B) the baseline cohort C) the cohorts after removing missing region and deprivation data, and D) the
